# Comparative Diagnostic Accuracy of Magnetic Resonance Elastography and Diffusion-Weighted Imaging in Differentiating Benign and Malignant Focal Liver Lesions: A Systematic Review and Meta-Analysis

**DOI:** 10.64898/2025.12.11.25342122

**Authors:** Amir Hassankhani, Parya Valizadeh, Payam Jannatdoust, Melika Amoukhteh, Abbas Mohammadi, Ali Gholamrezanezhad, Ali Haq

**Author notes:** Correspondence: Amir Hassankhani, MD.

## Abstract

**Background:** Accurate differentiation of benign and malignant focal liver lesions (FLLs) is essential for clinical decision-making. Magnetic resonance elastography (MRE) and diffusion-weighted imaging (DWI) are advanced MRI techniques used for noninvasive lesion characterization, but their comparative diagnostic performance has not been definitively established.

**Objective:** To systematically compare the diagnostic accuracy of MRE and DWI for distinguishing benign from malignant FLLs.

**Methods:** A systematic review and meta-analysis were conducted following PRISMA guidelines. PubMed, Embase, and Scopus were searched through July 2025 for studies directly comparing MRE and DWI in the same patient cohorts with focal liver lesions, using histopathology or validated imaging follow-up as the reference standard. Sensitivity, specificity, and area under the curve (AUC) were pooled using bivariate random-effects models, with paired analysis to compare modalities.

**Results:** 219 patients with 284 focal liver lesions were analyzed. MRE demonstrated higher pooled sensitivity (93.8%, 95% CI: 85.6–97.5) and specificity (89.9%, 95% CI: 74.6–96.4) than DWI (sensitivity 86.2%, 95% CI: 80.5–90.5; specificity 83.4%, 95% CI: 74.3–89.8). MRE also had a higher AUC (0.97 vs. 0.88). Likelihood ratio analysis indicated MRE’s stronger ability to both confirm and exclude malignancy. Paired meta-analysis confirmed a statistically significant increase in sensitivity for MRE (relative sensitivity 1.09; p = 0.018), with no significant difference in specificity.

**Conclusion:** MRE demonstrates superior sensitivity and overall diagnostic accuracy compared to DWI for differentiating benign and malignant FLLs. Further large-scale prospective studies are needed to confirm these results and determine optimal cutoff values to guide clinical decision-making.

## Introduction

Focal liver lesions (FLLs) are a common finding on cross-sectional abdominal imaging and often present a diagnostic challenge in clinical practice (1, 2). Although many FLLs—particularly in non-cirrhotic patients—are benign, distinguishing benign from malignant lesions at initial detection is critical because management, prognosis, and the need for further workup (including biopsy or oncologic staging) depend fundamentally on lesion etiology (3, 4). Consequently, noninvasive imaging plays a central role in lesion characterization and patient triage (4, 5).

Magnetic resonance imaging (MRI) is widely regarded as the most comprehensive modality for liver lesion evaluation due to its excellent soft-tissue contrast and ability to combine morphologic with physiologic and functional sequences (6, 7). Conventional contrast-enhanced multiphasic MRI remains a mainstay for lesion characterization; however, contrast administration may be contraindicated or undesirable in some patients, and atypical enhancement patterns can still lead to diagnostic uncertainty (8, 9). This has increased interest in non-contrast functional MRI techniques that probe tissue microstructure and mechanical properties as adjuncts or alternatives for noninvasive lesion assessment (9, 10).

Diffusion-weighted imaging (DWI) quantifies water motion within tissues and produces the apparent diffusion coefficient (ADC), which tends to be lower in highly cellular malignant tumors relative to many benign lesions (11, 12). Numerous studies have shown that ADC measurements can improve the discriminative performance of MRI for FLLs, although reported accuracy varies depending on b-value selection, acquisition technique, and lesion subtype (12–14). Limitations of DWI include overlap of ADC values between some benign and malignant lesions and susceptibility to motion and technical factors that affect reproducibility (15, 16).

Magnetic resonance elastography (MRE) offers a fundamentally different and complementary tissue descriptor: quantitative stiffness (17). By imaging mechanical wave propagation induced by an external driver, MRE estimates the viscoelastic properties of liver tissue and focal lesions (18). Early and recent studies suggest that many malignant liver tumors exhibit higher stiffness than surrounding parenchyma and benign lesions, making MRE a promising tool for lesion differentiation. While MRE is well established for staging diffuse liver fibrosis, its application to focal hepatic lesion characterization is less mature but rapidly evolving (17, 18).

Despite increasing research on both DWI and MRE for characterizing FLLs, direct comparisons between the two modalities are limited. To our knowledge, a rigorous synthesis of available evidence to quantify and compare the diagnostic performance of DWI and MRE in differentiating benign from malignant FLLs is still lacking.

Accordingly, this systematic review and meta-analysis aims to (1) identify and appraise studies directly comparing DWI and MRE for the characterization of FLLs, (2) pool diagnostic accuracy metrics (sensitivity, specificity, and area under the receiver-operating characteristic curve) where appropriate, and (3) compare the diagnostic performance of ADC-based DWI and stiffness-based MRE to determine their relative utility and potential complementary roles in clinical liver imaging. By synthesizing current evidence, our goal is to provide clinicians and researchers with clearer guidance on the strengths, limitations, and future directions for non-contrast MRI techniques in differentiating benign and malignant FLLs.

## Methods

### Study Design

This systematic review and meta-analysis were conducted following the Preferred Reporting Items for Systematic Reviews and Meta-Analyses (PRISMA) guidelines (19).

### Search Strategy

A comprehensive literature search was performed in PubMed, Embase, and Scopus from inception to June 28, 2025, to identify studies directly comparing MRE and DWI for differentiating benign and malignant FLLs. The search combined terms related to MRE, DWI, liver lesions, and diagnostic performance. The general search structure was as follows: (“magnetic resonance elastography” OR “MR elastography” OR MRE OR “MRI elastography”) AND (“diffusion-weighted imaging” OR “diffusion weighted imaging” OR DWI OR “diffusion MRI”) AND (“liver lesion*” OR “hepatic lesion*” OR “focal liver lesion*” OR “focal hepatic lesion*” OR tumor* OR mass* OR neoplasm*) AND (sensitivity OR specificity OR “diagnostic accuracy” OR “diagnostic performance” OR “receiver operating characteristic” OR ROC OR AUC OR detect* OR diagnos*).

Search terms were adapted appropriately for each database to maximize sensitivity and specificity. All retrieved records were imported into reference management software, where duplicates were removed prior to screening. Additionally, reference lists of included and relevant articles were manually screened to identify further eligible studies.

### Eligibility Criteria

Studies were included if they met the following criteria:

- Included human participants with suspected or confirmed FLLs;
- Performed both DWI and MRE within the same patient cohort;
- Provided a direct head-to-head comparison of the two modalities in differentiating benign from malignant lesions;
- Used histopathology or validated imaging follow-up as the reference standard;
- Reported or provided sufficient diagnostic accuracy data (true positives, false positives, true negatives, and false negatives) to allow calculation of sensitivity and specificity.

Exclusion criteria:

- Studies assessing only one modality (DWI or MRE) without direct comparison;
- Studies focusing exclusively on diffuse liver diseases without focal lesions;
- Publications lacking extractable diagnostic accuracy data;
- Case reports, small case series (<5 patients), conference abstracts without full data, reviews, editorials, and non-English studies without translation.

### Study Selection

Two reviewers independently screened titles and abstracts for eligibility. Full-text articles of potentially relevant studies were retrieved and assessed against inclusion criteria. Disagreements were resolved through discussion with a third reviewer.

### Data Extraction

Two reviewers independently extracted relevant data, including:

- Author, year, and country of publication;
- Study design and sample size;
- Patient demographics and inclusion criteria;
- Reference standards used for diagnosis;
- Technical details of DWI and MRE acquisition;
- Diagnostic accuracy metrics for each modality

Discrepancies were resolved by consensus.

### Quality Assessment

Methodological quality was independently assessed by two reviewers using both the Quality Assessment of Diagnostic Accuracy Studies-2 (QUADAS-2) tool (20) and the Quality Assessment of Diagnostic Accuracy Studies for Comparative Accuracy Studies (QUADAS-C) tool (21). QUADAS-2 evaluates risk of bias and applicability across four key domains: patient selection, index test, reference standard, and flow and timing. QUADAS-C extends this framework by incorporating an additional domain focused on the comparative nature of the studies, examining whether the head-to-head comparison of diagnostic tests was conducted fairly and without bias, including considerations such as test order, differential application of reference standards, and interpretation bias. Discrepancies in quality assessments between reviewers were resolved through discussion.

### Statistical Methods

The diagnostic test accuracy meta-analysis was performed using contingency data (true positives, true negatives, false positives, false negatives) extracted from each study. The comparative test accuracy meta-analysis was conducted in two steps:

- *Step 1:* A bivariate random-effects model, as described by Reitsma et al. (22), served as the primary analytical approach. Summary Receiver Operating Characteristic (SROC) curves were generated, with study weights proportional to their contribution in a univariate random-effects model of the diagnostic odds ratio (DOR). The area under the SROC curve (AUC) and corresponding 95% confidence intervals were calculated using 2000-sample parametric bootstrapping from the fitted bivariate model (23). The analysis was stratified by imaging technique (DWI and MRE) to enable direct comparison. Meta-regression based on the Reitsma bivariate model was then used to compare diagnostic accuracy between modalities. Between-study heterogeneity was quantified with the I² statistic (95% CI), with I² values above 50% indicating substantial heterogeneity (24). Due to the small number of included studies, no formal subgroup or meta-regression analyses were performed to explore heterogeneity sources. Diagnostic utility was further assessed using likelihood ratio scattergrams; positive likelihood ratios (LR s) >10 indicated strong rule-in value, and negative likelihood ratios (LR s) <0.1 indicated strong rule-out value (25). Pooled LR and LR values were calculated for each modality. All Step 1 analyses were conducted in R (version 4.2.1) using the packages “mada,” “meta” (26), “metafor” (27), and “dmetatools.“
- *Step 2:* A comparative, paired diagnostic test accuracy meta-analysis was conducted in Stata (StataCorp LLC, College Station, TX) using the metadta command (28). A bivariate random-effects logistic model was fit with test type (DWI vs MRE) as a meta-regression covariate applied to both logit-sensitivity and logit-false-positive rate. The paired structure (both modalities evaluated within the same studies) was handled using the comparative option in metadta (28), which modifies the linear predictor to provide valid comparative inference. From this model, pooled relative sensitivity and relative specificity (DWI vs MRE) were reported, and forest plots of relative accuracy were generated.

## Results

### Study Selection and Screening Process

An initial search using a predefined strategy yielded 377 records. After removing duplicates, 206 unique articles underwent title and abstract screening, resulting in the exclusion of 195 studies. The full texts of the remaining 11 articles were thoroughly evaluated, with 8 excluded for not meeting the study objectives. Ultimately, 3 articles met the inclusion criteria and were included in the analysis. The study selection process and eligibility criteria are summarized in a PRISMA-compliant flow diagram (Figure 1).

**Figure 1.**
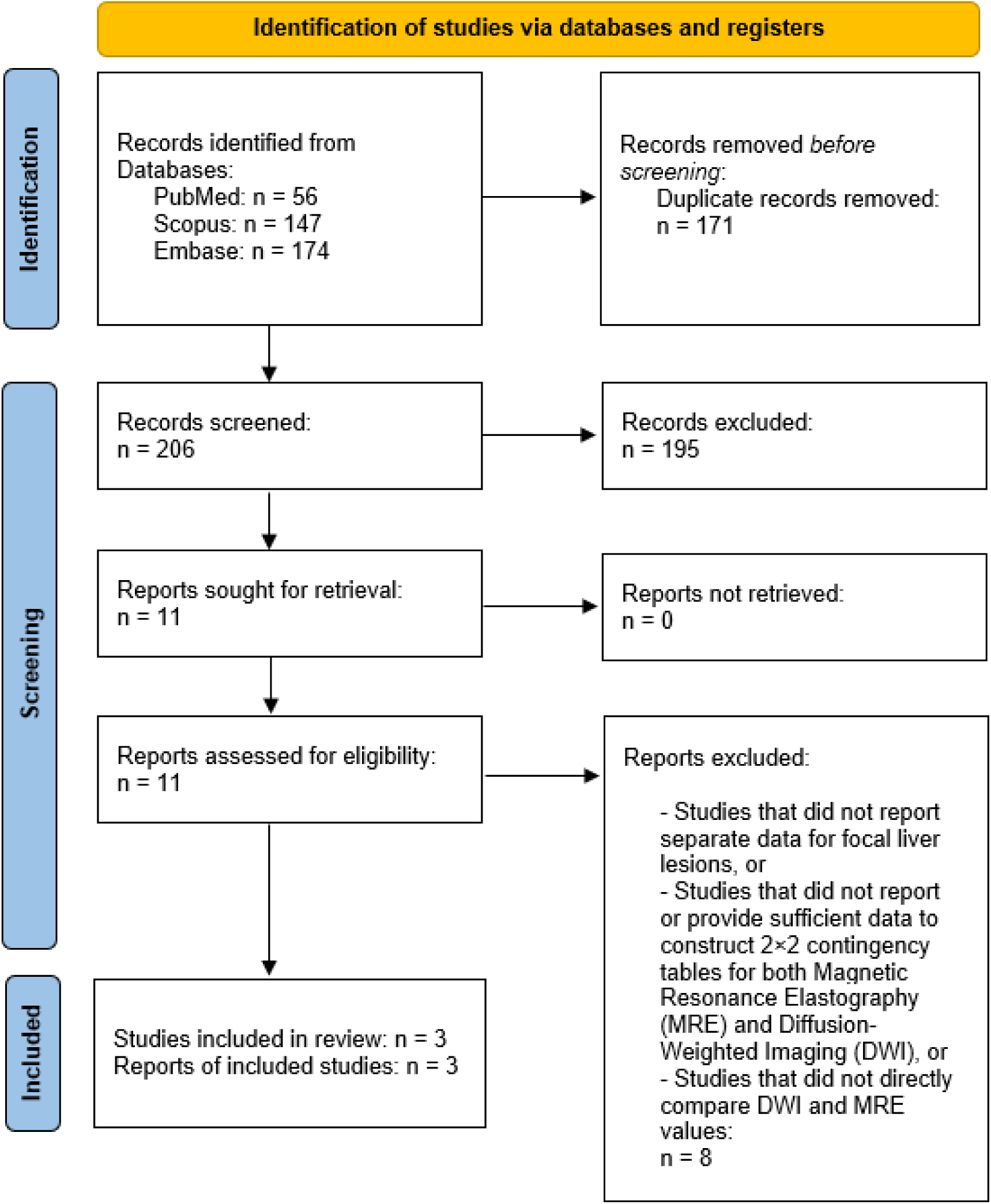
PRISMA 2020 flow diagram summarizing the study selection process for inclusion in the meta-analysis

### Study and Patient Characteristics

Table 1 summarizes the characteristics of the three included cross-sectional studies, conducted between 2016 and 2025, evaluating DWI and MRE for differentiating benign from malignant FLLs. Across all studies, a total of 219 patients were included, with 93 benign and 191 malignant lesions. Sample sizes ranged from 50 to 90 patients, with mean ages between 44.8 and 54.3 years. The proportion of malignant lesions was generally higher across studies, with common benign lesions including hemangiomas, focal nodular hyperplasia (FNH), and hepatocellular adenomas, and malignant lesions comprising hepatocellular carcinoma (HCC), metastases, cholangiocarcinoma, and in one study, lymphoma.

**Table 1.**
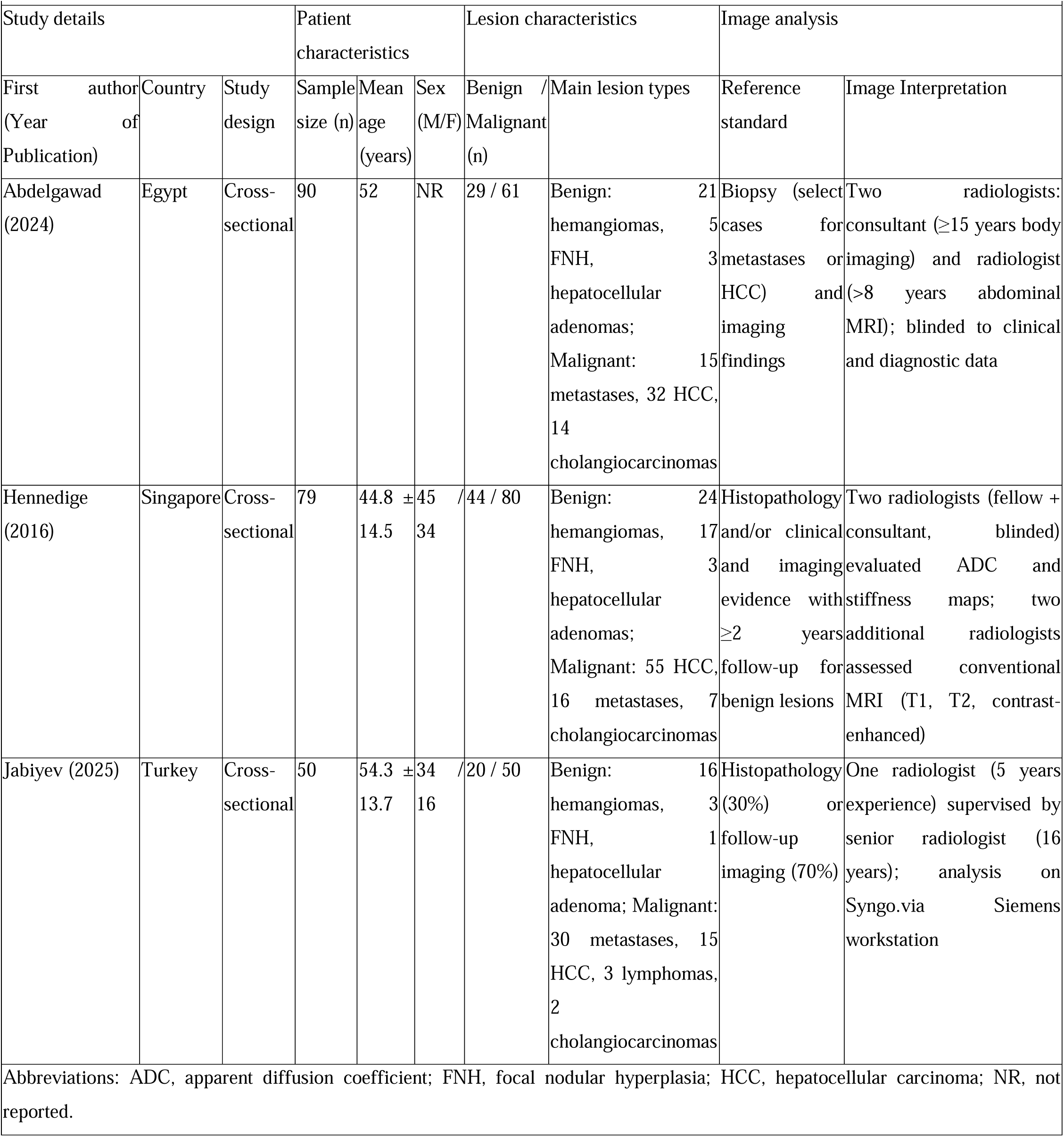
Characteristics of Included Studies Comparing DWI and MRE for Focal Liver Lesion Differentiation.

All studies used 1.5 T MRI scanners from different vendors, with DWI sequences varying in b-values and acquisition parameters, and MRE sequences based on modified gradient-echo techniques. Reference standards included histopathology alone or combined with long-term imaging follow-up. Image interpretation was performed by radiologists with varying experience levels, blinded to clinical information in all cases. Table 2 summarizes diagnostic accuracy metrics and optimal cut-off values for DWI and MRE in differentiating benign from malignant FLLs.

**Table 2.**
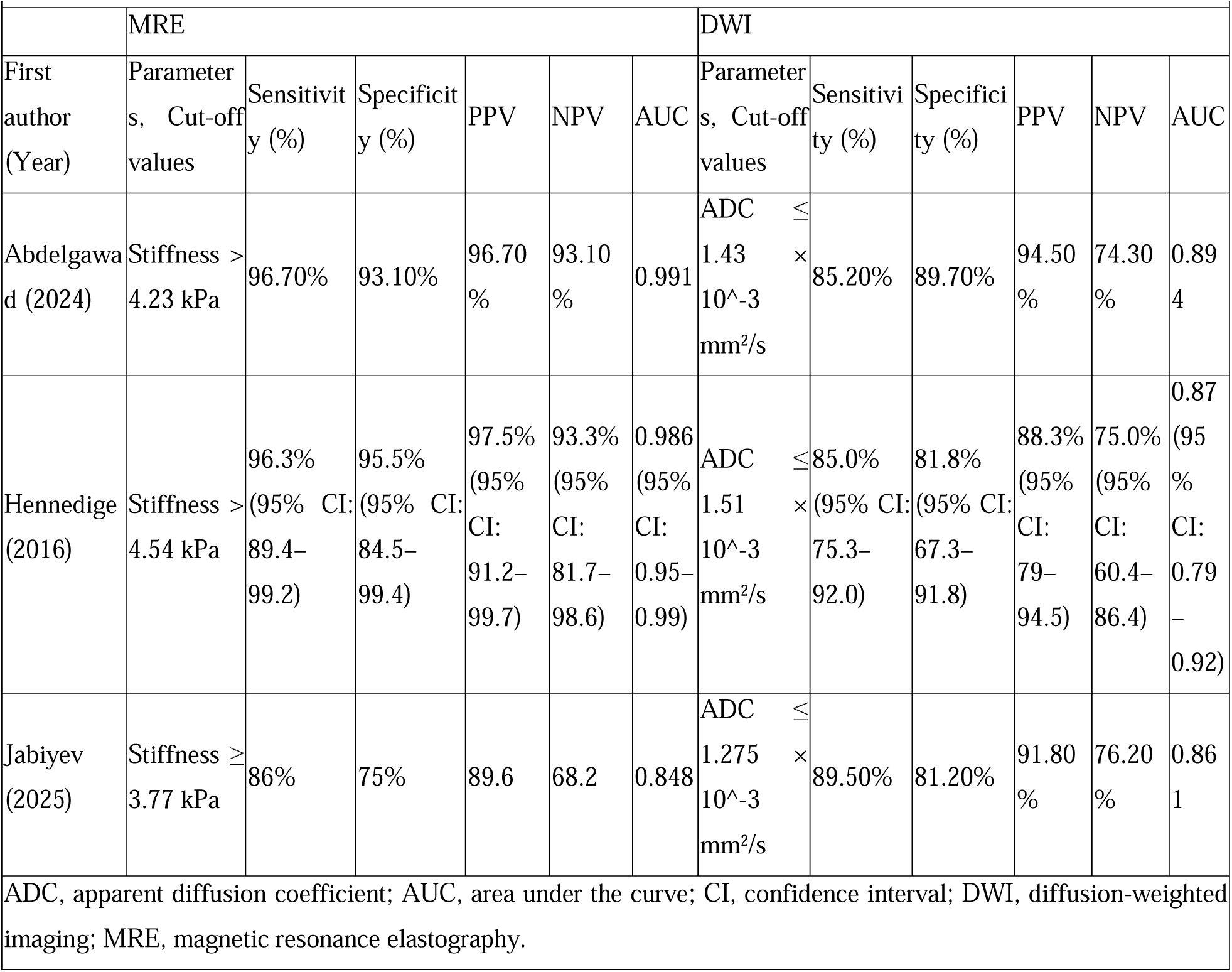
Diagnostic Performance of DWI and MRE for Differentiating Focal Liver Lesions in Included Studies.

|  | MRE |  |  |  |  |  | DWI |  |  |  |  |  |  |
| --- | --- | --- | --- | --- | --- | --- | --- | --- | --- | --- | --- | --- | --- |
| First author (Year) | Parameters, Cut-off values | Sensitivity (%) | Specificity (%) | PPV | NPV | AUC | Parameters, Cut-off values | Sensitivity (%) | Specificity (%) | PPV | NPV | AUC |  |
| Abdelgawad (2024) | Stiffness > 4.23 kPa | 96.70% | 93.10% | 96.70% | 93.10% | 0.991 | ADC $\leq 1.43 \times 10^{-3}$ mm<sup>2</sup>/s | 85.20% | 89.70% | 94.50% | 74.30% | 0.894 | |
| Hennedige (2016) | Stiffness > 4.54 kPa | 96.3% (95% CI: 89.4–99.2) | 95.5% (95% CI: 84.5–99.4) | 97.5% (95% CI: 91.2–99.7) | 93.3% (95% CI: 81.7–98.6) | 0.986 (95% CI: 0.95–0.99) | ADC $\leq 1.51 \times 10^{-3}$ mm<sup>2</sup>/s | 85.0% (95% CI: 75.3–92.0) | 81.8% (95% CI: 67.3–91.8) | 88.3% (95% CI: 79–94.5) | 75.0% (95% CI: 60.4–86.4) | 0.87 (95% CI: 0.79–0.92) | |
| Jabiyev (2025) | Stiffness ≥ 3.77 kPa | 86% | 75% | 89.6 | 68.2 | 0.848 | ADC $\leq 1.275 \times 10^{-3}$ mm<sup>2</sup>/s | 89.50% | 81.20% | 91.80% | 76.20% | 0.861 | |
| ADC, apparent diffusion coefficient; AUC, area under the curve; CI, confidence interval; DWI, diffusion-weighted imaging; MRE, magnetic resonance elastography. |  |  |  |  |  |  |  |  |  |  |  |  |  |

### Meta-Analysis

#### Overall analysis using bivariate model

Based on the bivariate model for diagnostic test accuracy meta-analysis, DWI achieved a pooled sensitivity of 86.2% (95% CI: 80.5–90.5) and pooled specificity of 83.4% (95% CI: 74.3–89.8), with no considerable heterogeneity (I² = 0.0%). In contrast, MRE demonstrated a higher pooled sensitivity of 93.8% (95% CI: 85.6–97.5) and specificity of 89.9% (95% CI: 74.6–96.4), though with substantial heterogeneity (I² = 79.9%–82.1%). Despite the notably higher sensitivity for MRE, there was no statistically significant difference in diagnostic accuracy metrics between groups (p = 0.261) (Figure 2).

**Figure 2.**
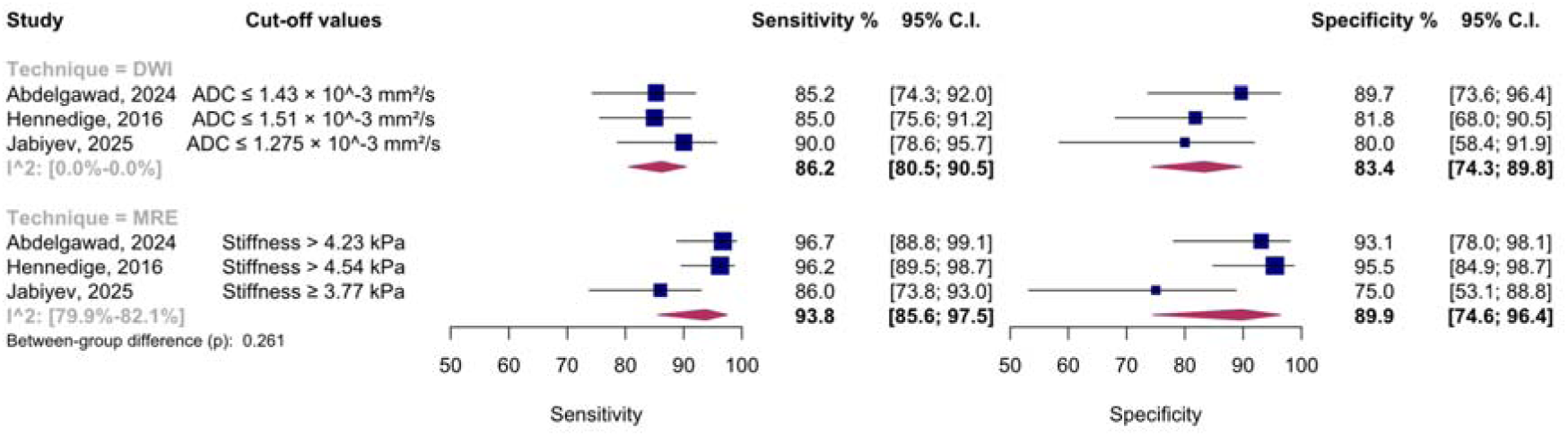
Forest plots showing pooled sensitivity and specificity (with 95% CIs) of DWI and MRE for differentiating benign from malignant focal liver lesions. **Abbreviations:** DWI, diffusion-weighted imaging; MRE, magnetic resonance elastography; ADC, apparent diffusion coefficient; CI, confidence interval; kPa, kilopascal.

SROC analysis with 2000-sample bootstrapping showed that DWI had a pooled AUC of 0.88 (95% CI: 0.82–0.94), while MRE achieved a higher pooled AUC of 0.97 (95% CI: 0.84–0.98) (Figure 3). The likelihood ratio matrix demonstrated that DWI fell within the quadrant indicating limited utility for either confirmation (LR□ < 10) or exclusion (LR□ > 0.1) of malignant liver lesions. In contrast, MRE, despite wide confidence intervals due to the limited number of studies, showed point estimates suggestive of potential utility for both confirmation (LR□ > 10) and exclusion (LR□ < 0.1) (Figure 4).

**Figure 3.**
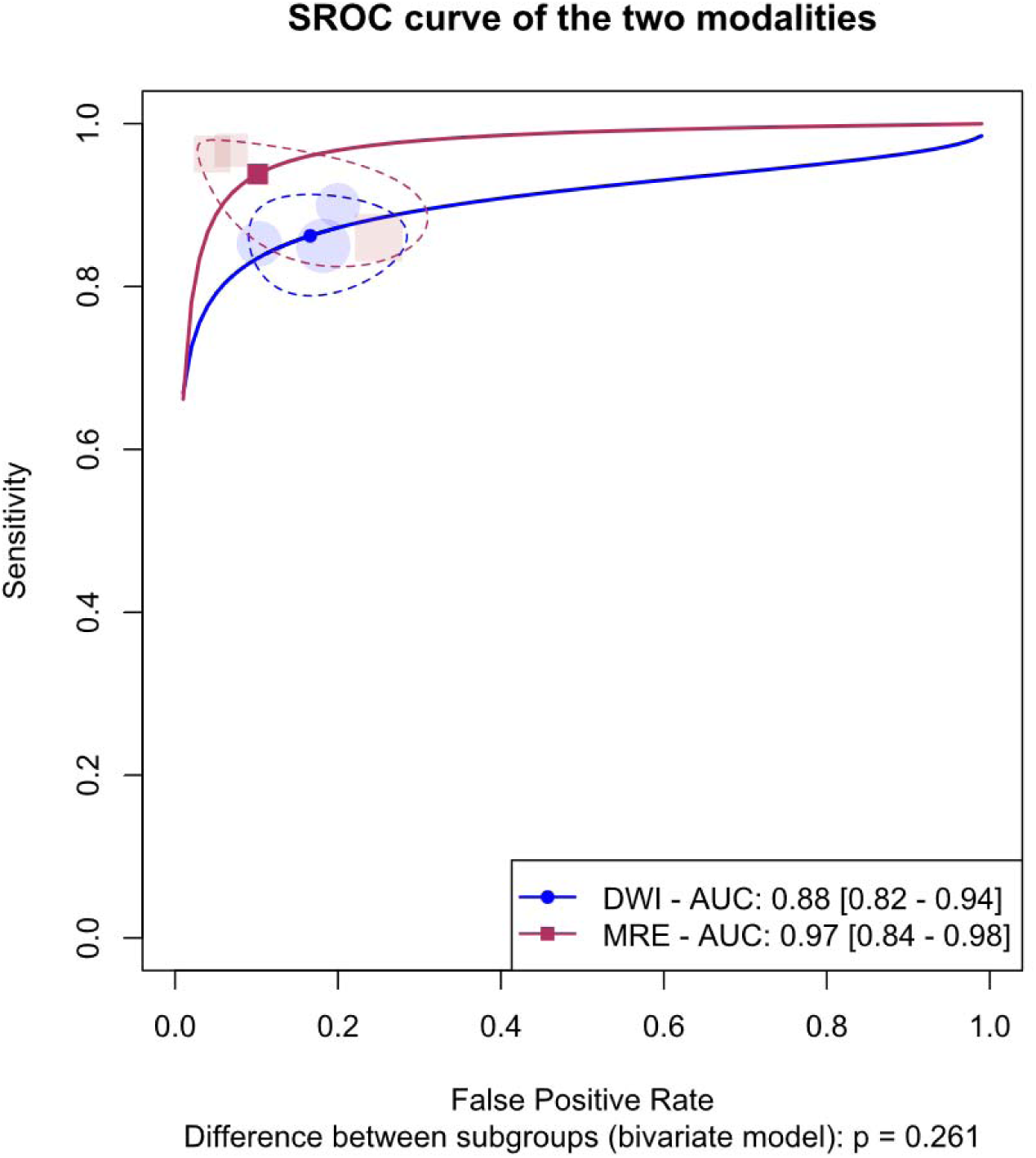
SROC curves for DWI and MRE in differentiating benign from malignant focal liver lesions. **Abbreviations:** DWI, diffusion-weighted imaging; MRE, magnetic resonance elastography; AUC, area under the curve; CI, confidence interval; SROC, summary receiver operating characteristic.

**Figure 4.**
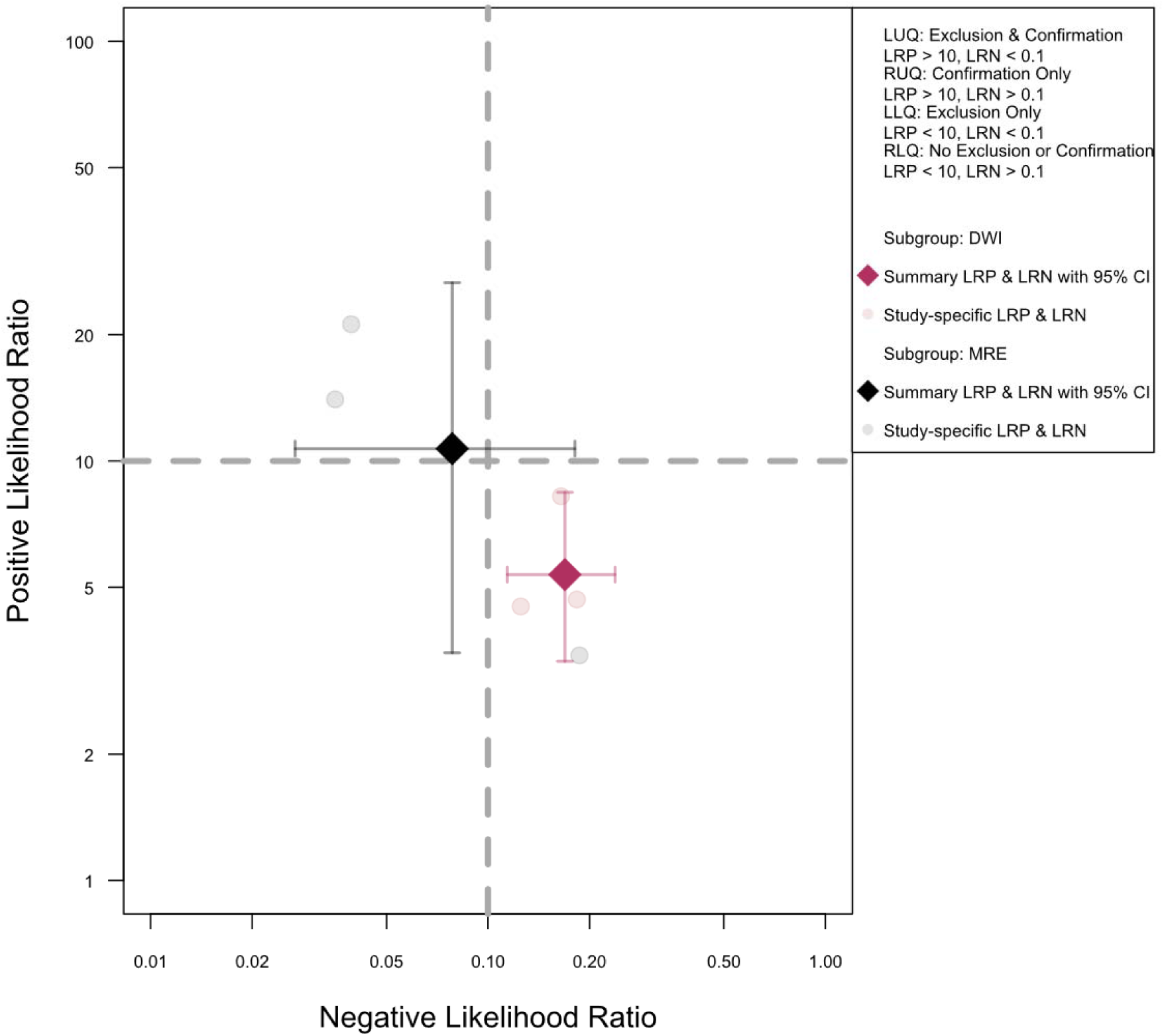
Likelihood ratio matrix plot for DWI and MRE in differentiating benign from malignant liver lesions. Each diamond represents the pooled LR□ and LR□ with 95% CIs for each technique, while grey circles indicate study-specific estimates. Dashed lines indicate conventional LR□ and LR□ thresholds for confirmation (>10) and exclusion (<0.1). **Abbreviations:** DWI, diffusion-weighted imaging; MRE, magnetic resonance elastography; LR□, positive likelihood ratio; LR□, negative likelihood ratio; CI, confidence interval; LUQ, left upper quadrant; RUQ, right upper quadrant; LLQ, left lower quadrant; RLQ, right lower quadrant.

Fagan nomograms indicated that for DWI (LR□ = 5.37, LR□ = 0.169), a positive test increased post-test probability of malignancy from pre-test probabilities of 25%, 50%, and 75% to 64.2%, 84.3%, and 94.2%, respectively, while a negative test reduced these probabilities to 5.3%, 14.5%, and 33.6% (Figure 5a). For MRE (LR□ = 10.7, LR□ = 0.078), corresponding post-test probabilities for a positive result were 78.1%, 91.45%, and 96.98%, while a negative result reduced them to 2.54%, 7.26%, and 19.02%, respectively (Figure 5b).

**Figure 5.**
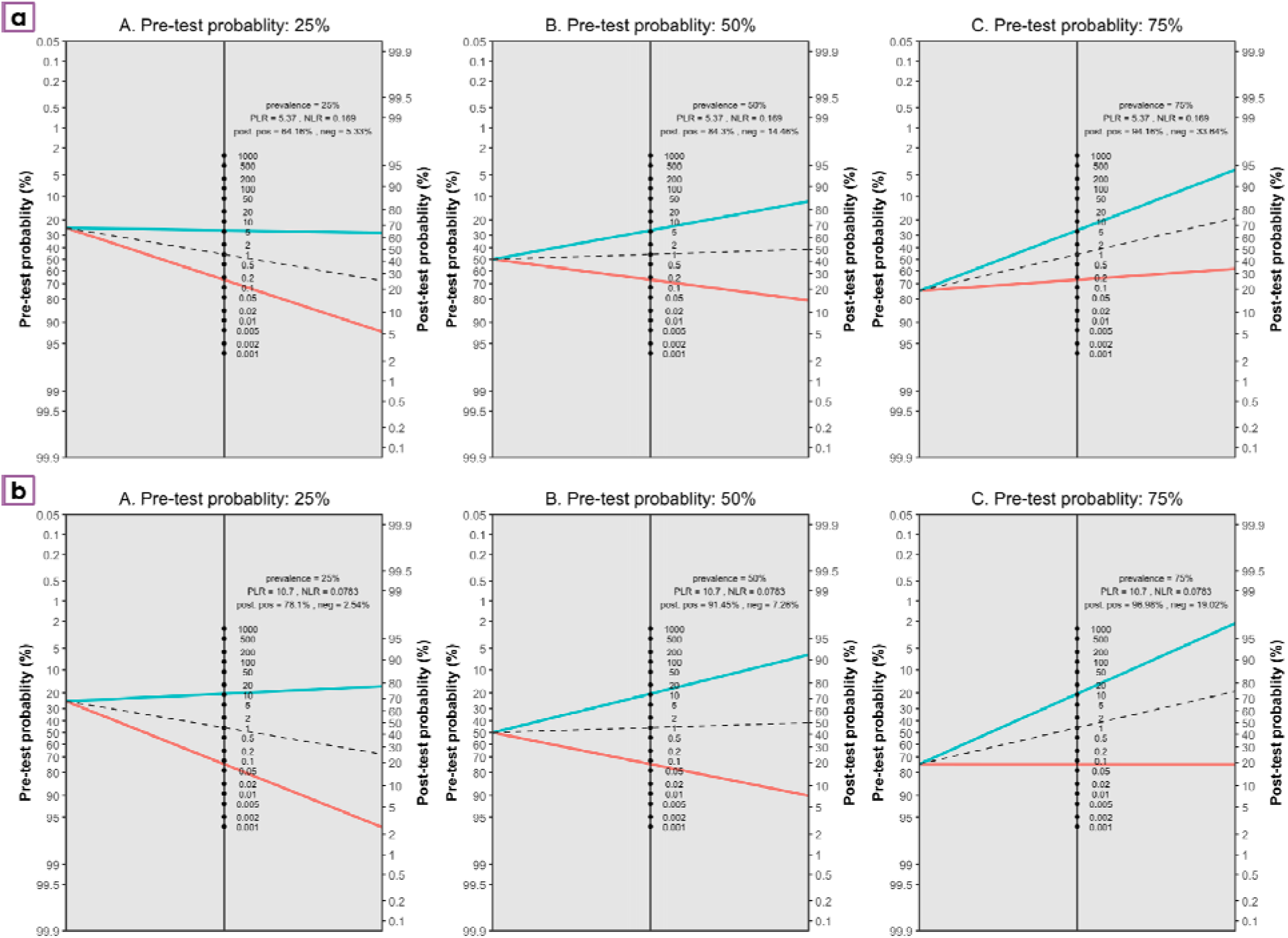
Fagan nomogram for DWI (a) and MRE (b), illustrating the effect of pre-test probabilities of 25% (A), 50% (B), and 75% (C) on post-test probabilities for malignancy in focal liver lesions. **Abbreviations:** DWI, diffusion-weighted imaging; PLR, positive likelihood ratio; NLR, negative likelihood ratio.

#### Comparative paired meta-analysis

The paired bivariate model, fitted with test type as a covariate, showed that MRE had a pooled relative sensitivity of 1.09 (95% CI: 1.01–1.16; p = 0.018) compared with DWI, indicating a modest but statistically significant increase in sensitivity. The pooled relative specificity was 1.08 (95% CI: 0.96–1.21; p = 0.197), showing no statistically significant difference in specificity between the two modalities. Comparative forest plots of relative sensitivity and specificity are presented in Figure 6.

**Figure 6.**
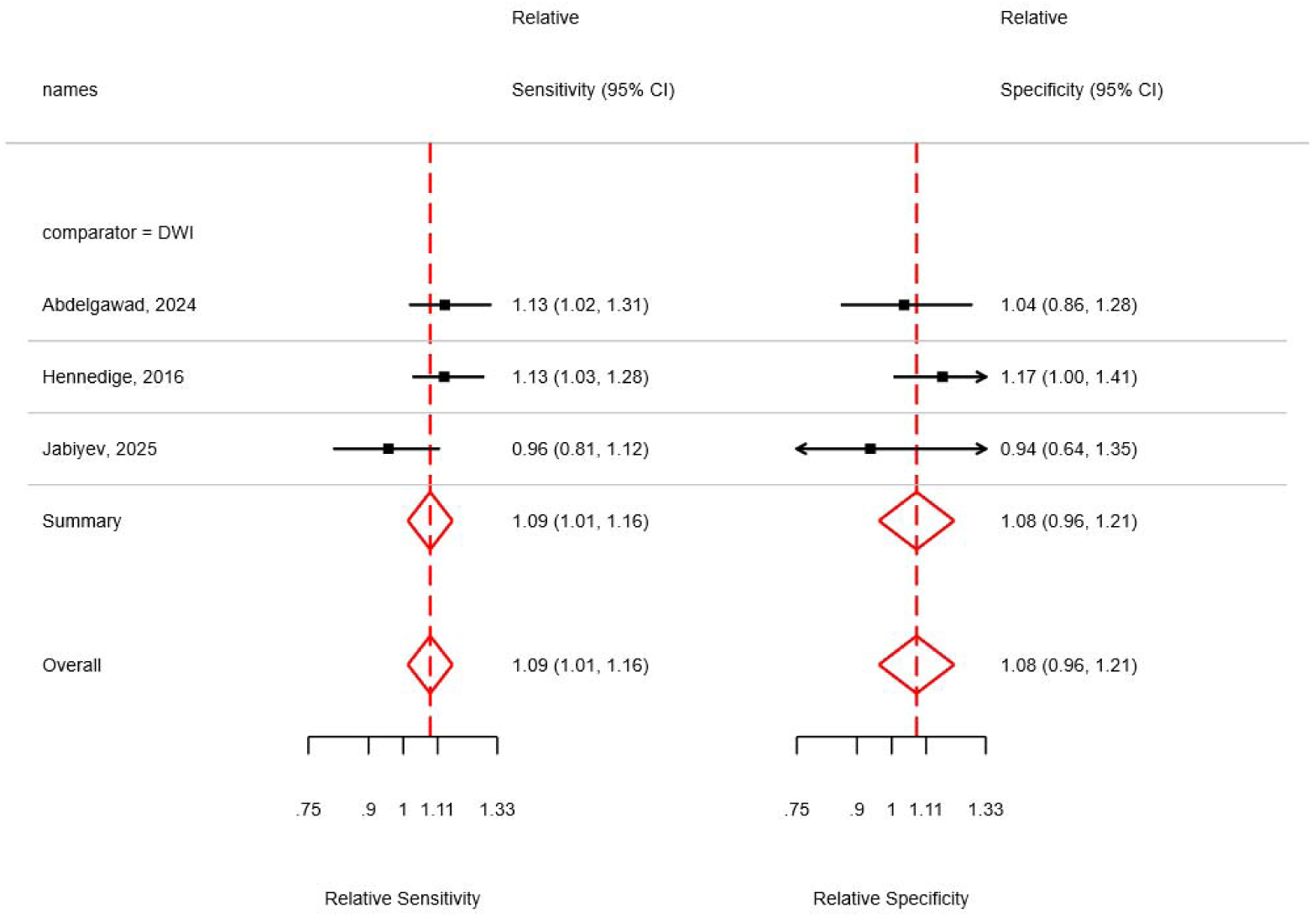
Paired comparative forest plots of relative sensitivity and relative specificity (with 95% confidence intervals) for MRE versus DWI in differentiating benign from malignant focal liver lesions. The comparator is DWI; effect sizes >1 favor MRE and <1 favor DWI. **Abbreviations:** DWI, diffusion□weighted imaging; MRE, magnetic resonance elastography; ADC, apparent diffusion coefficient; CI, confidence interval; kPa, kilopascal; RSens, relative sensitivity; RSpec, re

### Quality Assessment

Across the included studies, methodological quality was generally acceptable in most QUADAS-2 and QUADAS-C domains, with low applicability concerns and appropriate patient spectra. All studies used paired or fully comparable designs for MRE and DWI acquisition, with both tests performed within the same imaging session in most cases. However, a common limitation across all studies was the absence of pre-defined diagnostic thresholds, as cut-off values for both MRE and DWI. Additional concerns included limitations in the reference standard, particularly in Jabiyev et al. (29) and Hennedige et al. (30), where a substantial proportion of lesions were classified based on imaging follow-up rather than histopathology (Table 3).

**Table 3.**
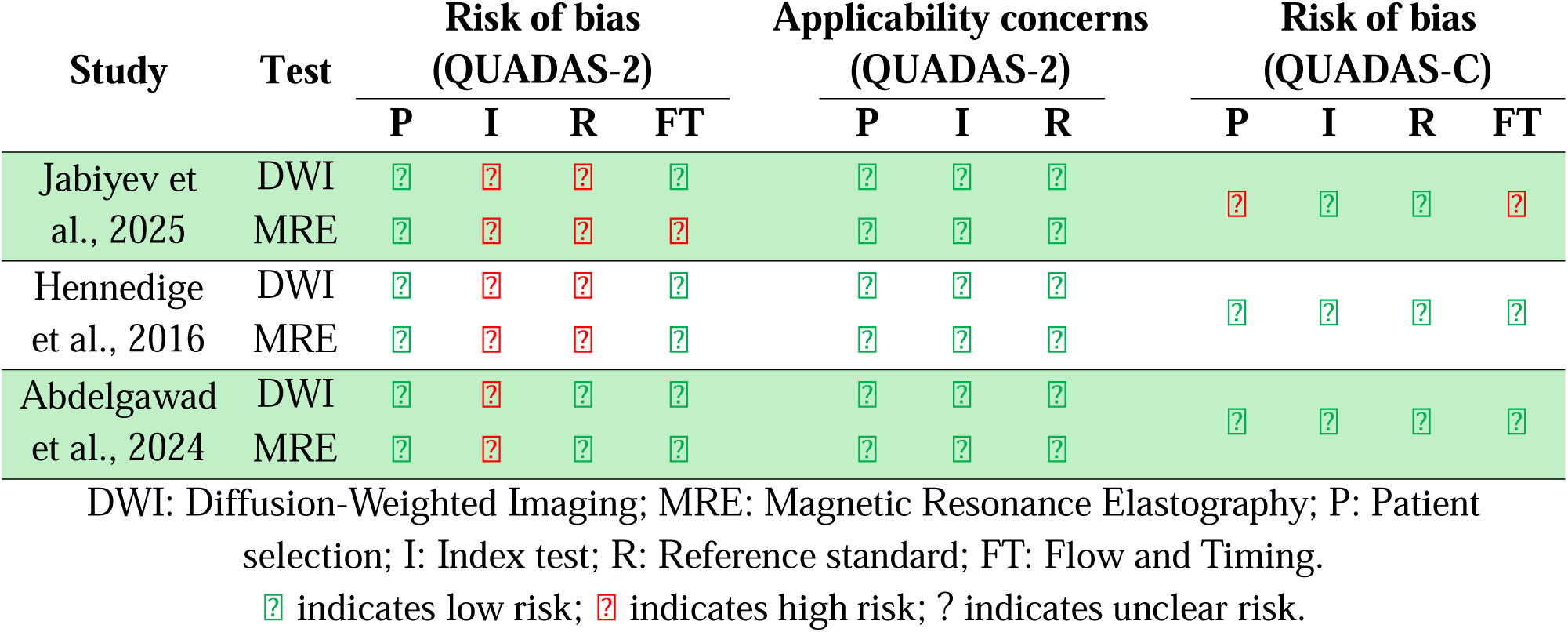
Risk of Bias Assessment of Included Studies Using QUADAS-2 and QUADAS-C.

## Discussion

This systematic review and meta-analysis is the first to directly compare DWI and MRE for differentiating benign and malignant FLLs. Despite the limited number of included studies, our findings offer valuable clinical insights that highlight MRE’s potential advantages in noninvasive liver lesion evaluation.

MRE demonstrated higher pooled sensitivity, specificity, and AUC than DWI, with likelihood ratio analyses supporting its superior ability to both confirm and exclude malignancy. The statistically significant increase in sensitivity suggests that MRE may more effectively reduce false-negative results, facilitating earlier diagnosis and timely treatment. This likely reflects MRE’s direct measurement of tissue stiffness—a hallmark of malignancy due to increased fibrosis and cellular density (31, 32)—whereas DWI assesses water diffusivity, which can be influenced by factors such as vascular perfusion, cellularity, and microcirculation, thereby limiting its specificity (30, 33).

Clinically, MRE’s ability to differentiate lesions using stiffness thresholds around 4 to 5 kPa with high accuracy is particularly valuable in complex contexts such as fibrotic or cirrhotic livers, where increased background stiffness can obscure other imaging findings (34, 35). This makes MRE especially useful for patients with chronic liver disease, who are at heightened risk for HCC and other malignancies. Moreover, multiple studies have demonstrated that malignant lesions—including cholangiocarcinoma and HCC—exhibit significantly higher stiffness than benign lesions, supporting MRE as a reliable biomarker of malignancy (31, 36, 37). In contrast, DWI—while valuable for assessing cellular density and diffusion restriction—suffers from overlapping ADC values between certain benign and malignant lesions, limiting specificity (38, 39). Additionally, DWI is susceptible to artifacts, low spatial resolution, and variability due to b-value selection, all of which affect reproducibility and interpretation (40, 41). These limitations underscore the potential complementary role of MRE when DWI results are equivocal.

The higher AUC observed for MRE further highlights its enhanced discriminatory power across diagnostic thresholds, suggesting more consistent performance and potentially greater diagnostic confidence than DWI. However, overlapping confidence intervals and significant heterogeneity in specificity warrant cautious interpretation. Larger prospective studies are needed to validate these findings and to establish standardized acquisition and interpretation protocols for MRE, which currently vary across platforms and institutions (29–31).

The likelihood ratio matrix further delineates clinical differences between the two modalities. While DWI provides useful diagnostic information, its limited ability to definitively rule in or out malignancy reduces its standalone utility. Conversely, MRE’s likelihood ratios suggest stronger potential as a decisive diagnostic test, which could reduce unnecessary biopsies and enable more timely treatment decisions. Nonetheless, wide confidence intervals reflect variability in current data and emphasize the need for more robust evidence. Additionally, Fagan nomogram analysis reinforces MRE’s superior clinical impact, demonstrating greater shifts in post-test probabilities for positive and negative results. MRE’s likelihood ratios exceed accepted thresholds for confidently ruling in or out malignancy, highlighting its potential to reduce diagnostic uncertainty and improve patient outcomes. While DWI enhances diagnostic certainty, its likelihood ratios are insufficient for definitive clinical decisions when used alone.

In summary, MRE appears to be a promising noninvasive imaging modality with superior diagnostic accuracy compared to DWI for characterizing liver lesions. However, the relatively small number of studies and patients contributed to heterogeneity in specificity estimates and limited subgroup analyses. Potential sources of this heterogeneity include variations in study populations, such as differing proportions of malignant and benign lesions, sample sizes, and mean patient age; technical differences, including MRI vendor, DWI b-values, and MRE sequence parameters; and variability in radiologist experience. Another key limitation is the lack of data stratified by baseline liver parenchymal conditions, such as fibrosis or steatosis, which may affect the accuracy of both MRE and DWI.

To build on this foundation, future research should focus on prospective trials with larger, diverse populations, standardize MRE acquisition and interpretation criteria to reduce variability, and explore combined use with other imaging modalities or clinical biomarkers to refine diagnostic algorithms. Studies should also assess how underlying liver parenchymal conditions influence diagnostic performance and evaluate cost-effectiveness and patient outcomes to support the clinical integration of MRE in liver lesion assessment.

## Conclusion

This meta-analysis demonstrates that MRE has significantly higher sensitivity and better overall diagnostic accuracy than DWI for distinguishing benign from malignant FLLs, with comparable specificity. MRE’s superior likelihood ratios indicate greater clinical utility for confirming or ruling out malignancy. Large-scale prospective studies are needed to validate these results and to develop standardized imaging protocols that optimize clinical decision-making and patient outcomes. Additionally, such studies should assess how baseline liver parenchymal conditions affect the diagnostic performance of both MRE and DWI.

## Data Availability

All data produced in the present study are available upon reasonable request to the corresponding author.

## References

1. Reizine E, Mulé S, Luciani A. Focal benign liver lesions and their diagnostic pitfalls. Radiol Clin North Am. 2022;60:755–773. 10.1016/j.rcl.2022.05.005

2. Wei Y, Yang M, Zhang M, et al. Focal liver lesion diagnosis with deep learning and multistage CT imaging. Nat Commun. 2024;15:7040. 10.1038/s41467-024-51260-6

3. Möller K, Safai Zadeh E, Görg C, et al. Prevalence of benign focal liver lesions and non-hepatocellular carcinoma malignant lesions in liver cirrhosis. Z Gastroenterol. 2023;61:526–535. 10.1055/a-1890-5818

4. James A. Focal lesions of the liver: imaging appearances and management. Br J Hosp Med (Lond). 2020;81:1–22. 10.12968/hmed.2019.0395

5. Wu H, Liang Y, Jiang X, et al. Meta-analysis of intravoxel incoherent motion magnetic resonance imaging in differentiating focal lesions of the liver. Medicine (Baltimore). 2018;97:e12071. 10.1097/MD.0000000000012071

6. Chavhan GB, Farras Roca L, Coblentz AC. Liver magnetic resonance imaging: how we do it. Pediatr Radiol. 2022;52:167–176. 10.1007/s00247-021-05053-4

7. Junhao L, Hongxia Z, Jiajun G, et al. Hepatic epithelioid angiomyolipoma: magnetic resonance imaging characteristics. Abdom Radiol (NY). 2023;48:913–924. 10.1007/s00261-023-03818-z

8. Bröker MEE, Taimr P, de Vries M, et al. Performance of contrast-enhanced sonography versus MRI with a liver-specific contrast agent for diagnosis of hepatocellular adenoma and focal nodular hyperplasia. AJR Am J Roentgenol. 2020;214:81–89. 10.2214/AJR.19.21251

9. Banerjee R, Pavlides M, Tunnicliffe EM, et al. Multiparametric magnetic resonance for the non-invasive diagnosis of liver disease. J Hepatol. 2014;60:69–77. 10.1016/j.jhep.2013.09.002

10. Yin M, Glaser KJ, Manduca A, et al. Distinguishing between hepatic inflammation and fibrosis with MR elastography. Radiology. 2017;284:694–705. 10.1148/radiol.2017160622

11. Shin MK, Song JS, Hwang SB, et al. Liver fibrosis assessment with diffusion-weighted imaging: value of liver apparent diffusion coefficient normalization using the spleen as a reference organ. Diagnostics (Basel). 2019;9:107. 10.3390/diagnostics9030107

12. Miyata T, Beppu T, Kuramoto K, et al. Hepatic sclerosed hemangioma with special attention to diffusion-weighted magnetic resonance imaging. Surg Case Rep. 2018;4:3. 10.1186/s40792-017-0414-z

13. Surov A, Eger KI, Potratz J, et al. Apparent diffusion coefficient correlates with different histopathological features in several intrahepatic tumors. Eur Radiol. 2023;33:5955–5964. 10.1007/s00330-023-09788-6

14. Chen J, Wu Z, Zhang Z, et al. Apparent diffusion coefficient and tissue stiffness are associated with different tumor microenvironment features of hepatocellular carcinoma. Eur Radiol. 2024;34:6980–6991. 10.1007/s00330-024-10743-2

15. Peng J, Li JJ, Li J, et al. Could ADC values be a promising diagnostic criterion for differentiating malignant and benign hepatic lesions in Asian populations: a meta-analysis. Medicine (Baltimore). 2016;95:e5470. 10.1097/MD.0000000000015470

16. Saleh GA, Razek AAKA, El-Serougy LG, et al. The value of the apparent diffusion coefficient in the Liver Imaging Reporting and Data System (LI-RADS) version 2018. Pol J Radiol. 2022;87:e43–e50. 10.5114/pjr.2022.113193

17. Liang J, Ampuero J, Castell J, et al. Clinical application of magnetic resonance elastography in hepatocellular carcinoma: from diagnosis to prognosis. Ann Hepatol. 2023;28:100889. 10.1016/j.aohep.2022.100889

18. Selvaraj EA, Mózes FE, Jayaswal ANA, et al. Diagnostic accuracy of elastography and magnetic resonance imaging in patients with NAFLD: a systematic review and meta-analysis. J Hepatol. 2021;75:770–785. 10.1016/j.jhep.2021.04.044

19. Moher D, Liberati A, Tetzlaff J, et al; PRISMA Group. Preferred reporting items for systematic reviews and meta-analyses: the PRISMA statement. PLoS Med. 2009;6:e1000097. 10.1371/journal.pmed.1000097

20. Whiting PF, Rutjes AW, Westwood ME, et al; QUADAS-2 Group. QUADAS-2: a revised tool for the quality assessment of diagnostic accuracy studies. Ann Intern Med. 2011;155:529–536. 10.7326/0003-4819-155-8-201110180-00009

21. Yang B, Mallett S, Takwoingi Y, et al; QUADAS-C Group; Bossuyt PMM, Brazzelli MG, et al. QUADAS-C: a tool for assessing risk of bias in comparative diagnostic accuracy studies. Ann Intern Med. 2021;174:1592–1599. 10.7326/M21-2234

22. Reitsma JB, Glas AS, Rutjes AWS, et al. Bivariate analysis of sensitivity and specificity produces informative summary measures in diagnostic reviews. J Clin Epidemiol. 2005;58:982–990. 10.1016/j.jclinepi.2005.02.022

23. Noma H, Matsushima Y, Ishii R. Confidence interval for the AUC of SROC curve and some related methods using bootstrap for meta-analysis of diagnostic accuracy studies. Commun Stat Case Stud Data Anal Appl. 2021;7:344–358. 10.1080/23737484.2021.1894408

24. Holling H, Böhning W, Masoudi E, et al. Evaluation of a new version of I2 with emphasis on diagnostic problems. Commun Stat Simul Comput. 2020;49:942–972. 10.1080/03610918.2018.1489553

25. Zwinderman AH, Bossuyt PM. We should not pool diagnostic likelihood ratios in systematic reviews. Stat Med. 2008;27:687–697. 10.1002/sim.2992

26. Balduzzi S, Rücker G, Schwarzer G. How to perform a meta-analysis with R: a practical tutorial. Evid Based Ment Health. 2019;22:153–160. 10.1136/ebmental-2019-300117

27. Viechtbauer W. Conducting meta-analyses in R with the metafor package. J Stat Softw. 2010;36. 10.18637/jss.v036.i03

28. Nyaga VN, Arbyn M. metadta: a Stata command for meta-analysis and meta-regression of diagnostic test accuracy data. Arch Public Health. 2022;80:140. 10.1186/s13690-021-00747-5

29. Jabiyev A, Karçaaltıncaba M, Karaosmanoğlu AD, et al. Multiparametric magnetic resonance imaging, diffusion-weighted magnetic resonance imaging, and magnetic resonance elastography: differentiating benign and malignant liver lesions. Diagn Interv Radiol. 2025. 10.4274/dir.2025.253324

30. Hennedige TP, Hallinan JT, Leung FP, et al. Comparison of magnetic resonance elastography and diffusion-weighted imaging for differentiating benign and malignant liver lesions. Eur Radiol. 2016;26:398–406. 10.1007/s00330-015-3835-8

31. Liang J, Qiu B, Yin S, et al. Predictive value of liver stiffness measurement by magnetic resonance elastography for complications after liver resection: a systematic review and meta-analysis. Digestion. 2022;103:357–366. 10.1159/000525081

32. Kim B, Kim SS, Cho SW, et al. Liver stiffness in magnetic resonance elastography is prognostic for sorafenib-treated advanced hepatocellular carcinoma. Eur Radiol. 2021;31:2507–2517. 10.1007/s00330-020-07357-9

33. Obara M, Kwon J, Yoneyama M, et al. Technical advancements in abdominal diffusion-weighted imaging. Magn Reson Med Sci. 2023;22:191–208. 10.2463/mrms.rev.2022-0107

34. Welle CL, Olson MC, Reeder SB, Venkatesh SK. Magnetic resonance imaging of liver fibrosis, fat, and iron. Radiol Clin North Am. 2022;60:705–716. 10.1016/j.rcl.2022.04.003

35. Xiao G, Zhu S, Xiao X, et al. Comparison of laboratory tests, ultrasound, or magnetic resonance elastography to detect fibrosis in patients with nonalcoholic fatty liver disease: a meta-analysis. Hepatology. 2017;66:1486–1501. 10.1002/hep.29302

36. Wu L, Bi J, Liu L, Zeng Y. Magnetic resonance elastography can predict the development of hepatocellular carcinoma: a meta-analysis and systematic review. J Gastrointest Oncol. 2021;12:1215–1222. 10.21037/jgo-21-196

37. Gao S, Sun W, Zhang Y, et al. Correlation analysis of MR elastography and Ki-67 expression in intrahepatic cholangiocarcinoma. Insights Imaging. 2023;14:204. 10.1186/s13244-023-01559-7

38. Testa ML, Chojniak R, Sene LS, et al. Is DWI/ADC a useful tool in the characterization of focal hepatic lesions suspected of malignancy? PLoS One. 2014;9:e101944. 10.1371/journal.pone.0101944

39. Nalaini F, Shahbazi F, Mousavinezhad SM, et al. Diagnostic accuracy of apparent diffusion coefficient (ADC) value in differentiating malignant from benign solid liver lesions: a systematic review and meta-analysis. Br J Radiol. 2021;94:20210059. 10.1259/bjr.20210059

40. Xiong H, Zeng YL. Standard-b-value versus low-b-value diffusion-weighted imaging in hepatic lesion discrimination: a meta-analysis. J Comput Assist Tomogr. 2016;40:498–504. 10.1097/RCT.0000000000000377

41. Calistri L, Castellani A, Matteuzzi B, et al. Focal liver lesions classification and characterization: what value do DWI and ADC have? J Comput Assist Tomogr. 2016;40:701–708. 10.1097/RCT.0000000000000458

